# THE SPREAD OF INFECTIOUS DISEASES IN MIGRATION ROUTES BETWEEN CARAVANS AND RESIDENT COMMUNITIES: MODELLING YELLOW FEVER IN CENTRAL AMERICA

**DOI:** 10.1101/2024.02.27.24303456

**Authors:** Sabrina Soares Simon, Marcos Amaku, Eduardo Massad

## Abstract

**Objectives:** To investigate the impact of yellow fever (YF) vaccination in the context of an outbreak and its association with migration, by assessing the effects of herd immunity in endemic communities and caravans, and its potential spread to other subsequent at-risk, disease-free communities in the migratory route.

**Methods:** We use a SEIRV-SEI compartmental deterministic model for humans and vectors and numerical simulations applied to three coupled populations - caravans, endemic and disease-free communities. Two scenarios are evaluated with different vaccination coverage levels at endemic communities and in caravans.

**Results:** When communities endemic to YF are the source of migration, herd immunity is critical, but vaccinating caravans is by far the most significant intervention to protect migrants and disease-free communities upwards from the risk of YF introduction.

**Conclusion:** Preventing outbreaks of climate-sensitive infectious diseases in the context of migration must be a joint effort of common interest. Maintaining vaccination coverage at recommended levels combined with the continued effort to vaccinate people on the move is the only intervention that can prevent a health crisis. Therefore, vaccination against climate-sensitive diseases should be considered climate adaptation.

## INTRODUCTION

As the 2018 Yellow Fever (YF) epidemic took place in South America, the awareness of vaccination coverage in the region increased, but even before the COVID-19 pandemic, the levels of vaccination and herd immunity began to decrease again. As a result, the vaccination levels in 11 of the 13 American endemic countries were below 95%, and in 7 of those countries, it was less than 80%.^1^ At the same time as migration is becoming an unprecedented crisis, it is impossible not to see a connection between human movement and climate-sensitive infectious diseases that could easily follow the flow with a susceptible population.

However, a superficial interpretation of this assumption can stigmatize migrants as mere vectors of diseases to be blamed for such phenomena.^2-4^ In contrast, people on the move seeking better living conditions are the most vulnerable to infectious disease outbreaks. The reason for fleeing a home country is frequently associated with the exact causes of climate-sensitive, (re)emerging diseases: a changing climate that deteriorates living conditions and reshapes the geographic distribution of vectors and other components of infectious diseases. As the current migration crisis in Latin America is strongly related to the climate crisis, the connection between these elements becomes clearer and more relevant every day in discussions for decisions that will shape our future in Planetary and Global Health.^5^

As migration reached unprecedented figures in Latin America, caravans have become more frequent and as extensive as 7,500 people traveling by foot, with nearly 20% composed of children walking about 25 miles a day. One of the reasons for such large groups is to guarantee safety, but the death rate of people on the move is still high: About 300 people have died or gone missing while migrating yearly in Latin America since 2014.^6-9^

The humanitarian sector is rapidly changing and faces a range of ethical, political and practical challenges associated with the climate crisis. In this scenario, mathematical models can be a powerful tool to raise awareness on key points of interventions. By connecting the study of climate-sensitive (re)emerging diseases and migration, both phenomena strongly shaped by the changing climate, we explore the relationship between the potential spread of YF and caravans in Central America, where the disease is not endemic but has a high risk of being introduced.^10^ Our goal is to provide simple and applicable modeling tools to project scenarios, evaluate the impact of vaccine interventions in YF outbreaks in the context of migration, and raise discussions towards the reevaluations of paradigms regarding migrants’ rights and stigma. We do not aim to offer system analysis and mathematical theorems or propositions but to reflect on the numerical simulations developed based on the study case results.

## METHODS

We use a SEIRV-SEI deterministic compartmental model based on a classical Ross–Macdonald model composed by ordinary differentiating equations with three coupled populations: (1) the community as the population of origin endemic for YF, (2) the caravan, and (3) the disease-free community as the destination. Each system of equations is composed of the following compartments for humans: Susceptible (*S*_*h*_), Exposed (*E*_*h*_), Infected (*I*_*h*_), Removed (*R*_*h*_), and Vaccinated (*V*_*h*_). Also, for mosquitoes, the compartments are Susceptible (*S*_*m*_), Exposed (*E*_*m*_), and Infected (*I*_*m*_).

The same conditions were applied to the stochastic simulations to account for the role of randomness in the scenarios. While deterministic models are suitable for large populations and accurately depict the overall pattern of an epidemic as it progresses, stochastic analysis can provide valuable insights, particularly in the early stages of an outbreak when only a small number of individuals are infected. The stochastic epidemic wave tends to be lower than the deterministic output, primarily due to the probability of the outbreak’s extinction as a result of random events. For this reason, the outcomes of stochastic simulations are considered more similar to real-life events and can be used to support and validate the findings provided by deterministic models.

All simulations were performed in the R environment using the “*lsoda*” function of the deSolve package.^11^ The Partial Rank Correlation Coefficient analysis was carried out with the epiR package^12^, and a stochastic modeling analysis was performed with the Binomial tau-leap Method from the Gillespie SSA2 package set for 100 simulations.^13^

### Model Formulation

This model covers a context where a caravan is formed in contact with a source community where YF is endemic. After the caravan leaves, it comes into contact with one or multiple disease-free communities on the way. When a caravan passes through a community, they occupy the same space and become a single population from the point of view of vector-borne diseases. Therefore, during this contact, the existence of an infected human or mosquito in any group represents a risk for the entire population.

The number of infected individuals entering a new population is determined by the day when the population comes into contact with the previous group where an outbreak is happening. The day (*t*_*n*_) when the caravan is formed and leaves the community of origin is considered to be the day when it comes into contact with this resident population. The day when the caravan arrives at the next community, which is disease-free, is regarded as the day of contact between them. For the numerical simulations presented, it was considered that both contacts happened on the 10^th^ day of an outbreak in the previous population. This choice is arbitrary and can be set according to the case of interest.

### Equations

The systems of equations are similar among the three populations, except for the disease-induced mortality rate (α_*h*_) being part of the dynamics of the community endemic for YF, and an additional death rate (θ_*c*_) applied to the migrant caravan due to the risks of the journey.

#### Human populations

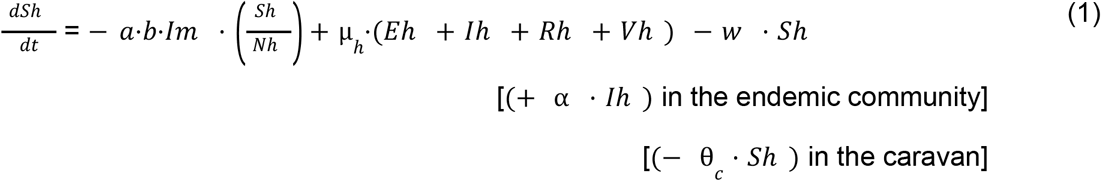

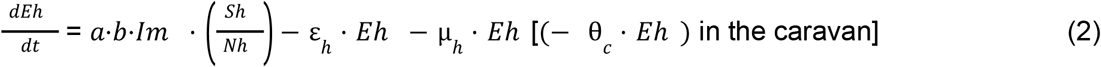

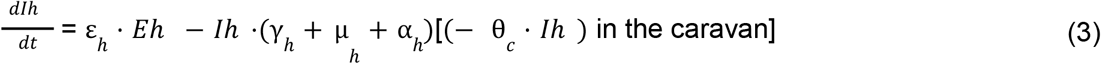

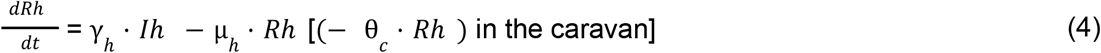

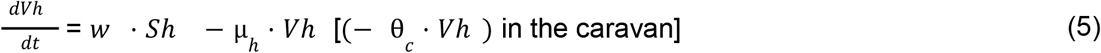

#### Mosquito population

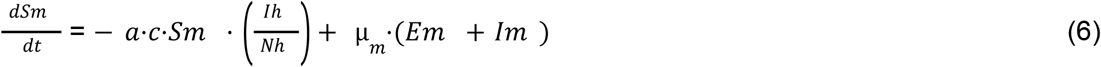

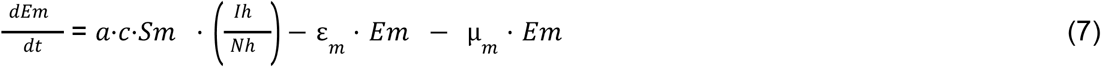

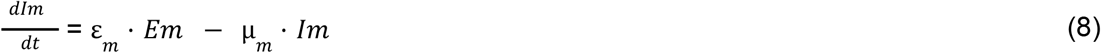

The primary reproduction number *R*_*0*_ was calculated by the well-known MacDonald equation for vector-borne diseases:

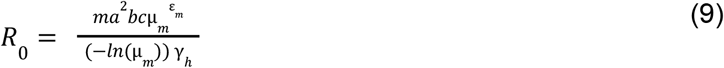

Being *m* the ratio of mosquitoes to humans 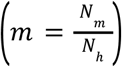. All the other parameters can be assessed in Table 1.

**Table 1:** Parameters and their biological meaning.

| *Parameter* | *Biological meaning* | *Daily rates* | *Source* |
| --- | --- | --- | --- |
| $a$ | Average biting rate | $0.33 \text{ day}^{-1}$ | 14 |
| $b$ | Fraction of infective bites | 0.25 | 15 |
| $c$ | Susceptibility of *Haemagogus* mosquito to the virus | 0.4 | 15 |
| $\mu_h$ | Natural mortality rate of humans | $3.77\text{e}^{-05} \text{ day}^{-1}$ | 16 |
| $\theta_c$ | Additional mortality rate of humans in transit | $1.917842\text{e}^{-05}$ | Newspapers and organization reports |
| $\mu_m$ | Natural mortality rate of *Haemagogus* mosquitoes | $0.0153 \text{ day}^{-1}$ | 17 |
| $\gamma_h$ | Human recovery rate | $0.1428 \text{ day}^{-1}$ | 18 |
| $\varepsilon_h$ | Latency rate in humans (intrinsic incubation period) | $0.167 \text{ day}^{-1}$ | 18 |
| $\varepsilon_m$ | Latency rate in mosquitoes (extrinsic incubation period) | $0.111 \text{ day}^{-1}$ | 18 |
| $\alpha_h$ | Disease-induced mortality rate | $8.0\text{e}^{-4} \text{ day}^{-1}$ | 14 |
| $w_1$ | Yearly vaccination rate in an endemic community | 0.1 to 0.95 | tested |
| $w_2$ | Yearly vaccination rate in the caravan/people on the move | 0 to 0.95 | tested |
| $w_3$ | Yearly vaccination rate in a disease-free community | 0.95 | tested |

### Initial conditions

The vaccination coverage in the endemic community before the outbreak represents its herd immunity. With the vaccination coverage of 95%, the proportion of susceptible individuals in this population at *t = 0* is given by

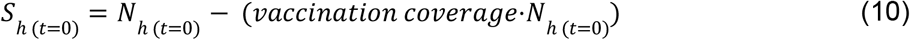

In the case of a community of one thousand people, there will be *S*_*h*(*t*=0)_ = 1000 − (0. 95 1000) = 1000 − 950 = 50 susceptible individuals at *t = 0*.

In the two other populations - caravan and disease-free - the number of susceptible individuals *S*_*h*_ equals the number of the entire population *N*_*h*_ when *t = 0*.

While an infected mosquito (*I*_*m*(*t*=0)_ = 1) with the YF virus (YFV) is responsible for entering the first population, the virus is introduced to the second and third populations by an infective human (*I* _*h* (*t*=0)_ = 1) each. **Assumptions**

- Vectors do not migrate. The vector population that corresponds to the caravans represents a continuum of space distribution along the migratory route;
- There is no climatic or environmental variation in time or space along the migratory route that could affect the dynamics or the components of the model;
- The caravan is formed only by susceptible individuals since it is difficult to estimate the herd immunity of such a heterogeneous group when many individuals may come from countries where the health system is weakened or close to collapse. In addition, a significant proportion of people on the move is made up of children under five years of age, which are probably naïve;
- All models are run for 365 days, although caravans usually do not last that long;
- The disease-free community is naïve since it is located outside the region of vaccination recommendation;
- The vaccination rate is considered only in the susceptible compartment. Therefore, exposed, infected, or already immune individuals may be vaccinated during vaccination campaigns, but that does not change the dynamics of the disease in the population.
- Vaccination is the only control measure taken into account.

## RESULTS

### 1. NUMERICAL SIMULATIONS

#### Scenario 1: Caravans are not vaccinated

Three vaccination coverages (or annual vaccination rates) are considered in the resident, endemic community: 0.1, 0.5, and 0.95. To isolate the effect of the vaccination in this community upon the subsequent populations, vaccination rates were fixed at 0.00 in the caravan and 0.95 in the disease-free community. It is important to highlight that vaccination campaigns started along with the YF outbreak and not prior. The three populations have the same size of 1000 people.

The parameters provided in Table 1 show that the effective reproduction number *R*_*0*_ for YF calculated with equation (9) is 6.83. This means that in order to keep the proportion of susceptible individuals (S/N) under the value of 1/*R*_*0*_, the vaccination coverage must reach above the minimum of 85.36% of the population.

Figure 1 shows that the effects of vaccination in the endemic community on the naive caravan are not so much in reducing the number of cases but mainly in delaying the process, allowing time for other interventions to take place.

**Fig. 1:**
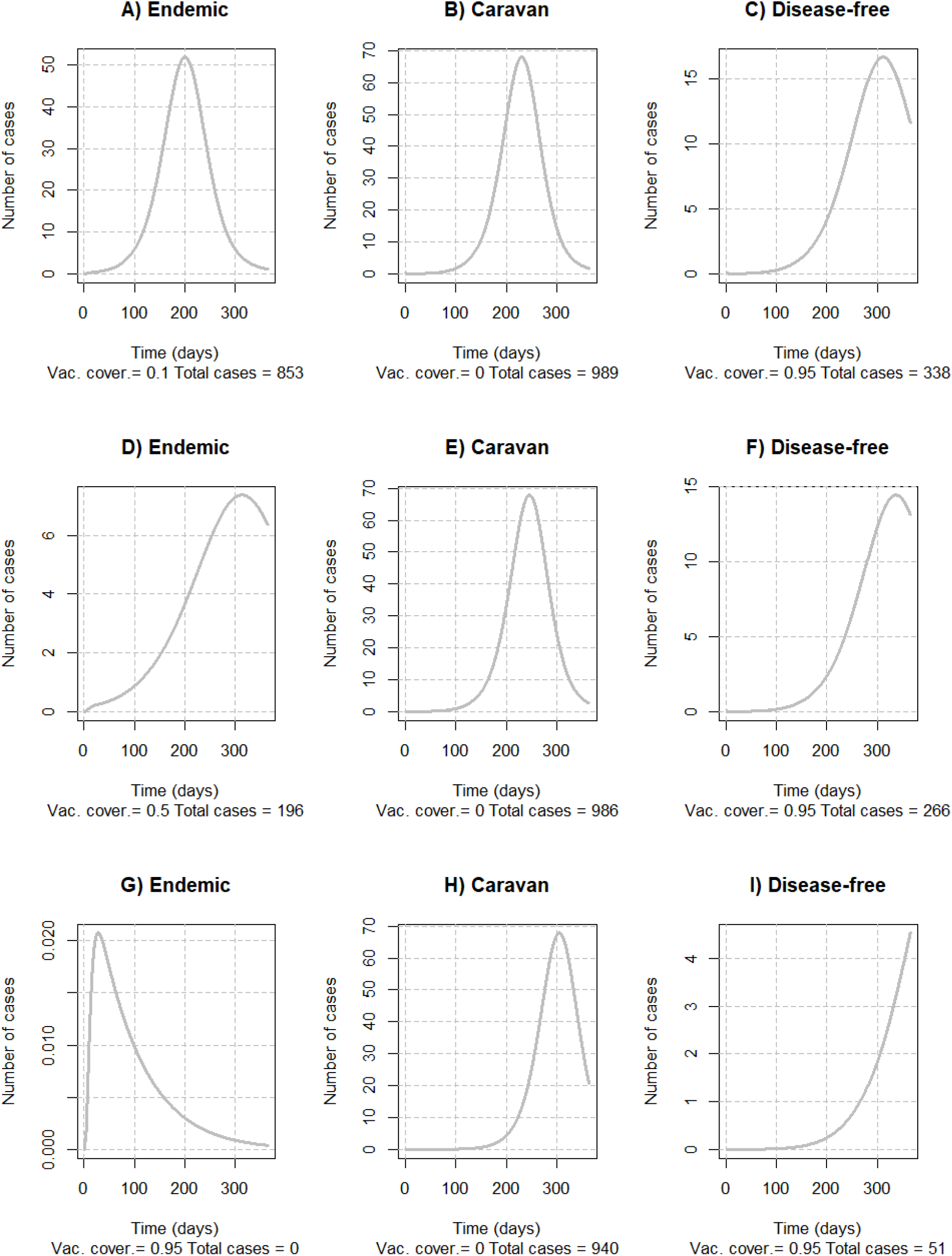
Effects of the YF vaccination of endemic communities over subsequent communities. A-C: Vaccination coverage in the endemic community is 10%; D-F: Vaccination coverage in the endemic community is 50%; and G-I: Vaccination coverage in the endemic community is 95%. Attention to the different y-axis.

#### Scenario 2: Migrant caravans are vaccinated

To isolate the effects of vaccination in the surrounding communities, the vaccination coverage of both resident endemic and disease-free communities was fixed at 0.95. That way, the impact of three different vaccination levels in the caravan (0.0, 0.5, and 0.95) is presented in Figure 2.

**Fig. 2:**
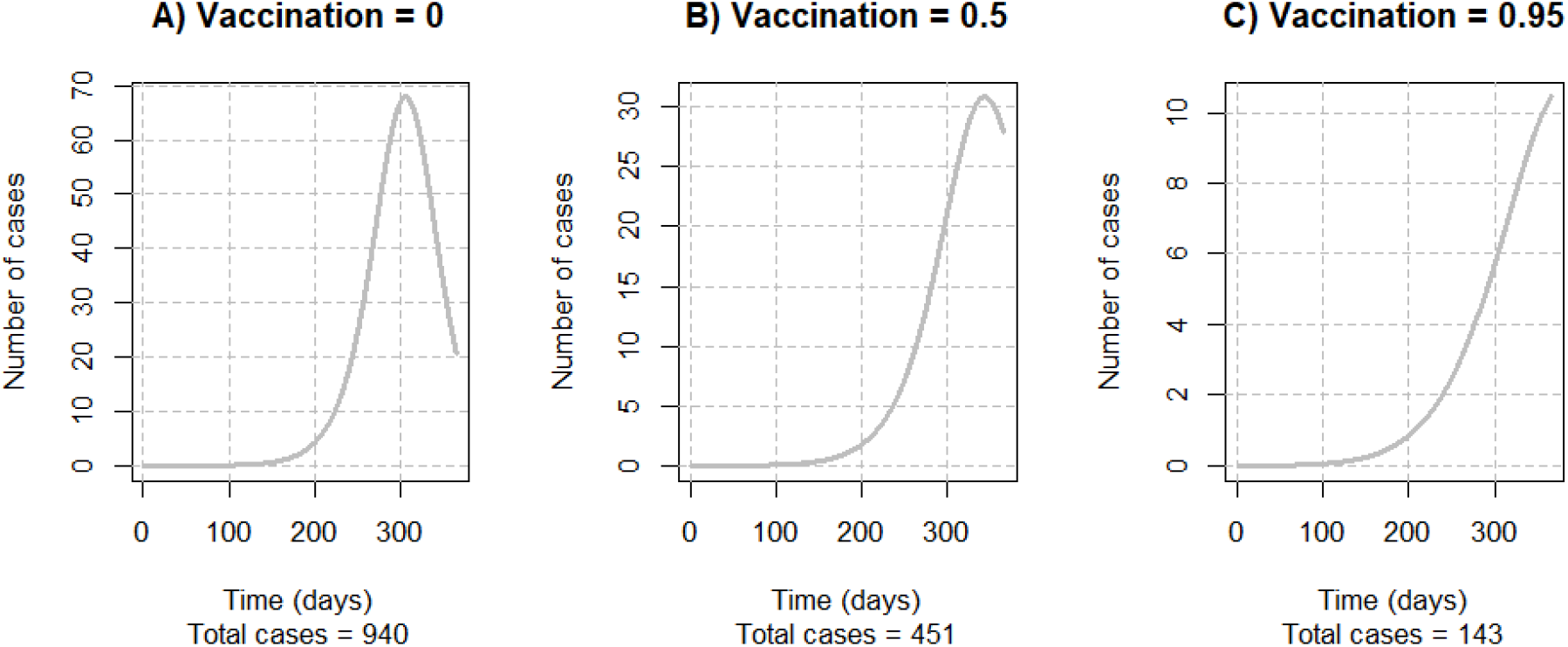
Effects of three vaccination rates on caravans (0%, 50%, and 95% a year) considering a vaccination coverage of 95% in all the surrounding communities.

Vaccinating people migrating in groups can reduce the number of cases and considerably delay the outbreak’s evolution, allowing new interventions to contribute even when the campaign begins simultaneously with the outbreak.

### 2. SENSITIVITY ANALYSIS

According to the Partial Rank Correlation Coefficient Analysis (Fig. 3), the most relevant parameters to the model are related to the biology of the vector that determines the infection rate: the biting rate *(a)*, the fraction of infective bites *(b)*, and vector susceptibility *(c)* and its natural mortality, in addition to the recovery rate in humans (γ_*h*_). Among the vaccination rates in the three populations *(w*_*1*_, *w*_*2*_, *w*_*3*_), the one related to the source community has the most powerful impact on the number of cases *(w*_*1*_ = 0.48), followed by the vaccination in the caravan, and lastly, at the disease-free community.

**Fig. 3:**
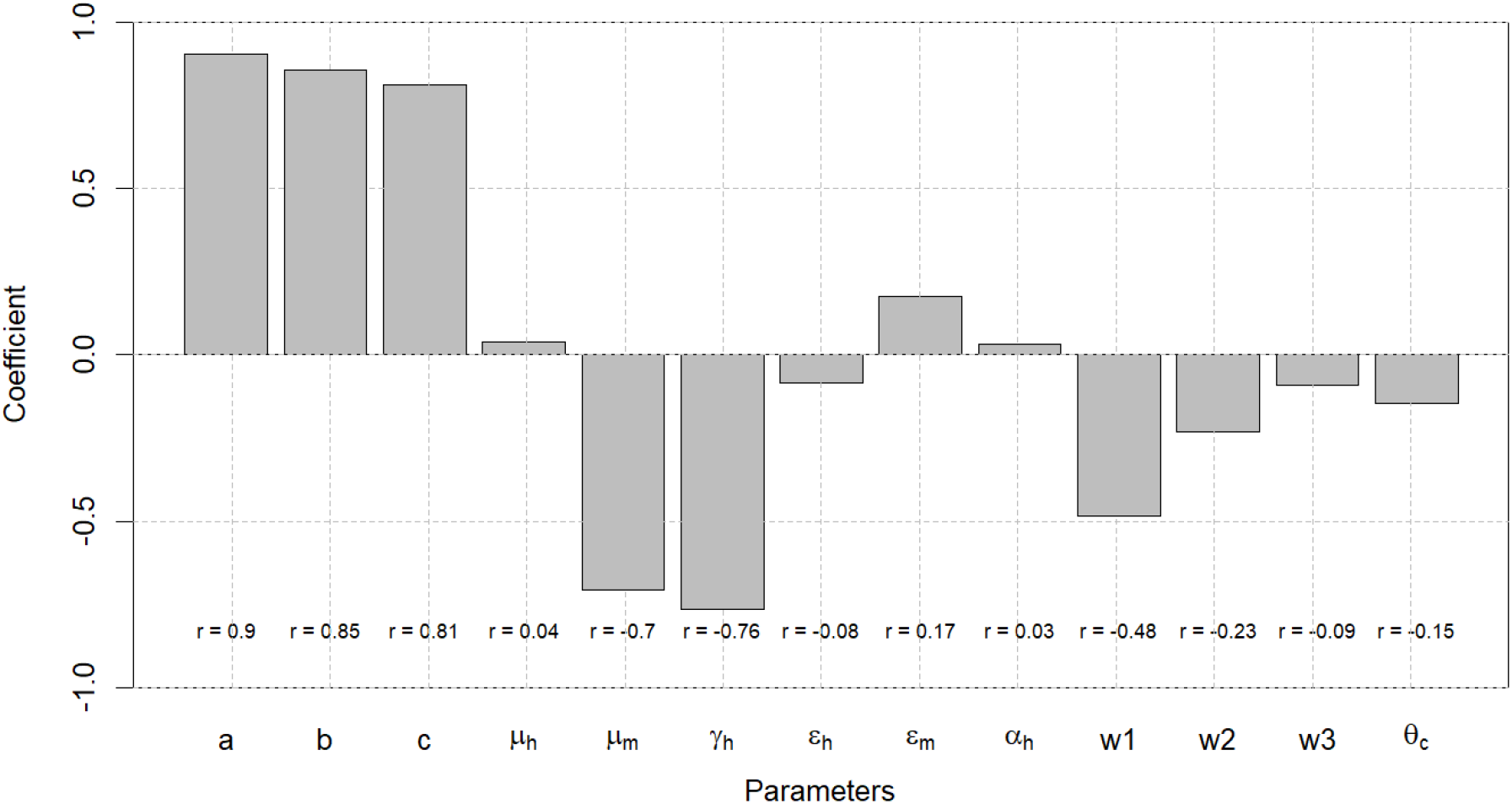
PRCCA for each of the parameters in the model, *w*_*1*_ the vaccination rate for the endemic community, while *w*_*2*_ corresponds to the caravan and *w*_*3*_ to the disease-free community.

### 3. STOCHASTIC MODELLING

The graphs in Figure 4 illustrate the diverse range of potential outcomes in terms of the duration and magnitude of the epidemic curve when comparing the best-case and worst-case scenarios to the outcomes provided by the deterministic model (black line). Despite this variability, both scenarios are corroborated by the projections given by the deterministic model. Lower vaccination coverage amplifies the impact of randomness on the outbreak, leading to a higher probability of disease extinction before an epidemic occurs, particularly when vaccination exceeds recommended levels (95% coverage). Each grey line in the background represents a stochastic simulation and thus, a potential outcome.

**Fig. 4:**
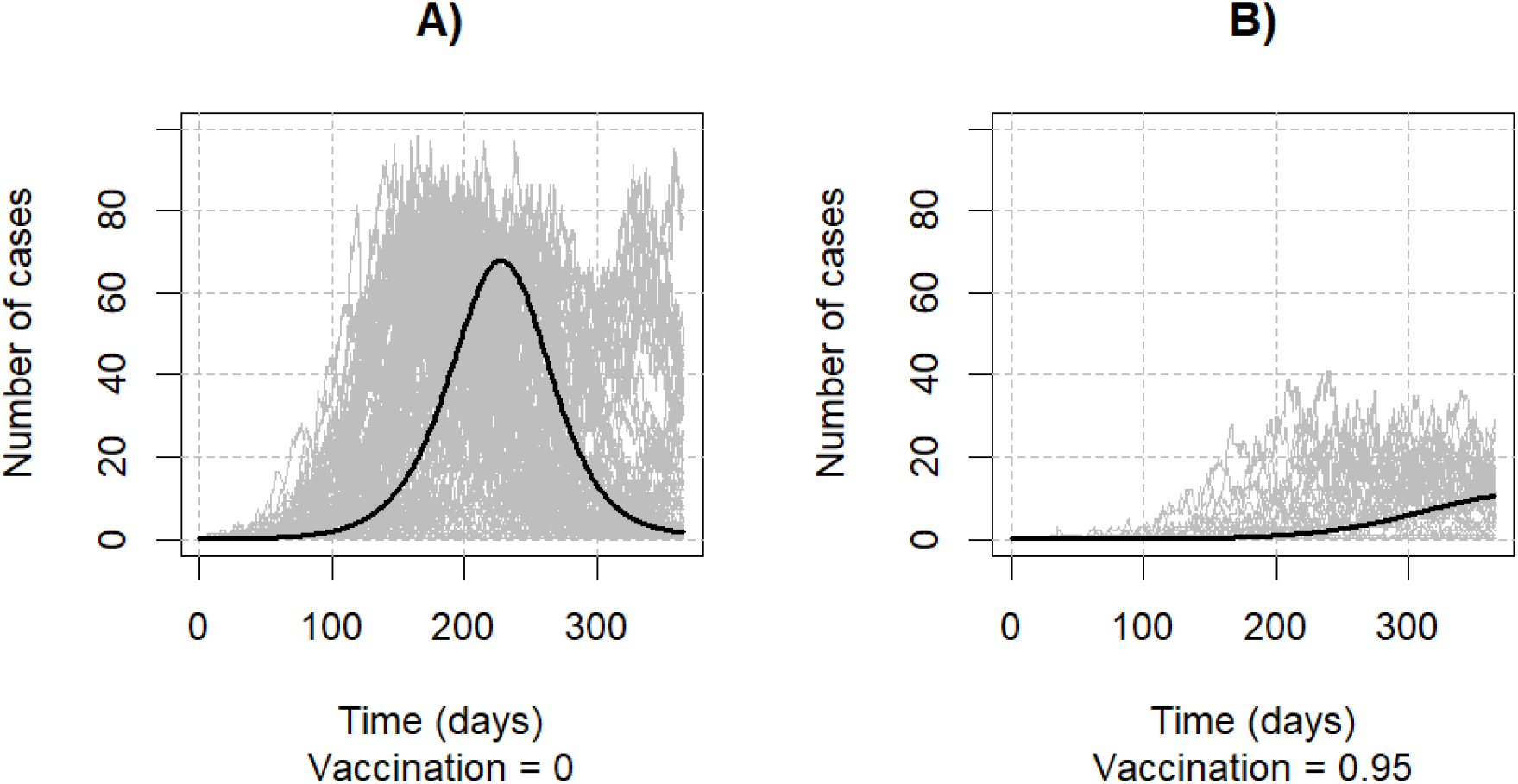
Stochastic simulation of the effects of A) vaccination coverage = 0.0% and B) 95% on the caravan, for a group of 1000 people. Grey lines represent the noise of 100 simulations, while the black line represents the projection given by the deterministic simulation presented earlier. Vaccination coverage of surrounding populations is fixed at 95%.

## DISCUSSION

This model shows that caravans, like any other group, can be infected by contacting a resident endemic community, not only the other way around. Resident endemic communities with low herd immunity offer a risk to caravans passing through, and this risk might be taken to other communities on the route without effective vaccination coverage. Of course, any vaccination level is better than none, but the higher the vaccination coverage in endemic communities, the more protected the subsequent populations will be. However, without vaccinating caravans, there will be no effective control of a potential outbreak that could spread to the routes or introduce the virus in other disease-free communities.

Even vaccinated, caravans are still the most vulnerable groups in all the scenarios presented. This is due to the lack of herd immunity and the day of contact with the source community and its epidemiological conditions: the smaller the number of infected individuals, the slower the *R*_*0*_, which will delay the next population in the sequence. When *R*_*0*_ increases in the first population, the later the contact, the greater the risk of introduction in the second population due to the higher proportion of infective individuals coming from the source population. Moreover, caravans face additional risks due to the risks of the journey itself, which can lead to an overestimation or underestimation of the death toll, and there is no additional birth rate to compensate for it.

The combined effects of vaccination efforts of the two resident communities are powerful, even considering that the disease-free community begins to vaccinate after the first case arises, showing how important it is to take vaccination campaigns as a joint effort among nations. The World Health Organization has raised this issue, since, in 2021, the 95% vaccination level was not achieved by 12 of the 13 endemic countries for YF in Latin America, while 8 of those reached less than 80%.^19^ However, the only intervention that could significantly protect caravans is vaccination, which can reduce the number of cases by 85% if the campaign starts simultaneously with the outbreak.

The main message is that having 95% of the population vaccinated before the outbreak is more valuable than 95% of the vaccination rate as the outbreak begins, but this is only mandatory for endemic countries. Consistently vaccinating caravans is the best way to protect communities ahead by giving them time to respond to a potential outbreak. However, despite other priorities, vaccination against climate-sensitive diseases has been left behind in the assistance of migrants and caravans.^20^ It has been found that migration does not cause outbreaks when the host population has sufficient herd immunity. In cases like this, small levels of migration can dilute the disease in both source and host populations.^21^

If migration alone were able to spread viruses, YFV would probably be already endemic in Central and North America. For this to occur, there must be a combination of environmental factors that climate change may promote shortly which can expand the geographic distribution of competent vectors northwards. If this happens, YFV will be endemic in most places of destination, but not due to migration. Migrants are infinitely more vulnerable to infections, while the responsibility for spreading climate-sensitive infectious diseases is the lack of consistent vaccination and a changing climate. Only a deliberate and consistent collective vaccination effort can act as a public health barrier, guaranteeing safe migration as a human right and a climate adaptation for migrants, residents, and endemic and disease-free communities.

The reason deterministic models often appear overly catastrophic compared to our perception of reality stems from the role of randomness. This is why simulations sometimes predict hundreds of cases, yet in reality, the disease frequently extinguishes before reaching epidemic levels. Stochastic simulations consider the possibility of disease extinction even under conditions conducive to an outbreak, particularly in its early phases. Nevertheless, it’s essential to emphasize that, with the conditions and parameters applied in our study, an epidemic of significant magnitude, as predicted, remains plausible, even in the presence of random events. The stochastic analysis supports the findings of deterministic simulations, which encompass a range of potential outcomes.

The stochastic simulation is an example of a technique that copes with an important shortcoming of deterministic models. In general, the main limitation of modeling studies is the simplistic assumption of reality, especially in such a complex scenario as the Latin American migration crisis. As the model grows in complexity, it can be a powerful tool, but it may become unable to make accurate projections due to the lack of data. For example, caravans have a very changeable composition, and taking into account the turnover by people daily entering and exiting the system would bring a layer of complexity that would probably not increase the accuracy of the projection. In addition, tracking caravans and estimating the contact with adjacent communities is a challenge yet to be arranged. Finally, the simulations presented are built on multiple arbitrary decisions, but while the amount and the quality of data increase, mathematical models can grow as a powerful tool for implementing impactful results. Still, as simple as they are, models like this can bring insightful discussions and support decision-making around challenging contexts.

## Data Availability

All data produced in the present work are contained in the manuscript.

## PUBLIC HEALTH IMPLICATIONS

Caravans represent a population on the move that connects both endemic and disease-free neighboring communities. We realize that maintaining vaccine coverage above recommended levels in endemic communities is crucial to prevent the YFV from being introduced into at-risk regions, acting as a filter against disease expansion into new areas. However, it is the massive vaccination of caravans that effectively protects adjacent communities along the way and, above all, can reduce the preventable suffering of people in migration - the most vulnerable components of the system. Once again, vaccination campaigns derived from international and cross-border cooperation strategically located are key to ensuring climate adaptation for migrating groups and surrounding communities, especially those where the disease is not yet circulating endemically.

## STATEMENT OF ETHICS REVIEW

This project did not involve human data or participants, therefore, per the guidelines of the Plataforma Brasil Institutional Review Board, IRB assessment was not necessary.

## REFERENCES

1. Pan American Health Organization. Yellow fever in the Region of the Americas: Vaccine reserve stockpile management - 26 May 2022 [Internet]. PAHO/WHO; 2022 May 26 [cited 2023 Jan 30]. Available from: https://www.paho.org/en/documents/yellow-fever-region-americas-vaccine-reserve-stockpile-management-26-may-2022

2. Tasker A, Braam D. Positioning zoonotic disease research in forced migration: A systematic literature review of theoretical frameworks and approaches. PLoS ONE. 2021;16(7):e0254746. doi: 10.1371/journal.pone.0254746.

3. Saeed F, Mihan R, Mousavi SZ, Reniers RL, Bateni FS, Alikhani R, et al. A Narrative Review of Stigma Related to Infectious Disease Outbreaks: What Can Be Learned in the Face of the Covid-19 Pandemic? Front Psychiatry. 2020 Dec 2;11:565919. doi: 10.3389/fpsyt.2020.565919.

4. Silva TM, Cade MV, Figueiras A. Impact of infectious disease epidemics on xenophobia: A systematic review. J Migr Health. 2022;5:100085. 10.1016/j.jmh.2022.100085.

5. Climate Change and Health: An Urgent New Frontier for Humanitarianism. 2018 [cited 2023 Feb 03]. Available from: https://www.researchgate.net/publication/329352488_Climate_Change_and_Health_an_urgent_new_frontier_for_humanitarianism.

6. The Republic | azcentral.com. For the migrant caravan, the true distance of the journey ahead begins to sink in [Internet]. 2018 [cited 2023 Feb 03]. Available from: https://www.azcentral.com/story/news/politics/border-issues/2018/10/28/migrant-caravan-journey-has-thousands-miles-go-united-states-border/1797645002/.

7. The Washington Post. One day and 28 miles with the migrant caravan in Mexico: Thirst, songs and a police showdown [Internet]. 2018 [cited 2023 Jun 30]. Available from: https://www.washingtonpost.com/world/the_americas/one-day-and-28-miles-with-the-migrants-in-mexico-thirst-songs-and-a-police-showdown/2018/10/29/7c763455-c2f3-4f46-8a8e-f2263901266b_story.html

8. OIM Oficina Regional para Centroamérica, Norteamérica y el Caribe. DTM - Reporte situacional 9 de flujos de personas migrantes en tránsito observadas [Internet]. [cited 2023 Jun 30]. Available from: https://rosanjose.iom.int/es/resources/dtm-reporte-situacional-9-de-flujos-de-personas-migrantes-en-transito-observadas

9. Missing migrants | Migration Data Portal [Internet]. The International Organization of Migration (IOM) ‘s Missing Migrants Project, 2022. [cited 2023 Jan 31]. Available from: https://www.migrationdataportal.org/themes/migrant-deaths-and-disappearances

10. Shearer FM, Longbottom J, Browne AJ, Pigott DM, Brady OJ, Kraemer MUG, et al. Existing and potential infection risk zones of yellow fever worldwide: a modelling analysis. Lancet Glob Heal. 2018 Mar;6(3):e270–8.

11. Soetaert K, Petzoldt T, Setzer RW. Solving Differential Equations in R : Package deSolve. J Stat Softw. 2010;33(9).

12. CRAN - Package epiR [Internet]. [cited 2023 Jan 30]. Available from: https://cran.r-project.org/web/packages/epiR/index.html

13. Cannoodt R, Saelens W, Deconinck L, Saeys Y. Spearheading future omics analyses using dyngen, a multi-modal simulator of single cells. Nat. Commun. 2021;12:3942.

14. Massad E, Amaku M, Coutinho FAB, Struchiner CJ, Lopez LF, Wilder-Smith A, et al. Estimating the size of Aedes aegypti populations from dengue incidence data: Implications for the risk of yellow fever outbreaks. Infect Dis Model. 2017 Nov;2(4):441–54.

15. Esteva L, Vargas C, Yang HM. A model for yellow fever with migration. Comput Math Methods. 2019 Nov 1;1(6):e1059.

16. The United Nations. World Population Prospects 2019: Highlights | Multimedia Library - United Nations Department of Economic and Social Affairs [Internet]. 2019 [cited 2022 Jul 21]. Available from: https://www.un.org/development/desa/publications/world-population-prospects-2019-highlights.html

17. Raimundo SM, Yang HM, Massad E. Modeling Vaccine Preventable Vector-Borne Infections: Yellow Fever as a Case Study. J Biol Syst. 2016;24(2):1–24.

18. Pan American Health Organization. Control of Yellow Fever: Field Guide - PAHO/WHO | Pan American Health Organization [Internet]. 2005 [cited 2022 Jul 26]. Available from: https://www.paho.org/en/documents/control-yellow-fever-field-guide

19. Epidemiological Alert: Yellow Fever - 31 August 2022 - PAHO/WHO | Pan American Health Organization [Internet]. [cited 2023 Jan 31]. Available from: https://www.paho.org/en/documents/epidemiological-alert-yellow-fever-31-august-2022

20. International Activity Report 2021. Medicin Sans Frontiere [Internet]. [cited 2023 Feb 03]. Available from: https://www.msf.org/international-activity-report-2021

21. Burton J, Billings L, Cummings DAT, Schwartz IB. Disease persistence in epidemiological models: the interplay between vaccination and migration. Math Biosci. 2012 Sep;239(1):91–6.

